# Unmet Needs in Acute Hepatic Porphyria Diagnosis: A Comparative Big Data Analysis of an AI-based Human-in-the-Loop Screening Versus Standard of Care

**DOI:** 10.1101/2025.03.28.25324843

**Authors:** Simon Lin, Georg Strebinger, Melanie Kaiser, Kathrin Blagec, Vinzenz Pilgram, Thomas Lutz, Jama Nateqi

## Abstract

**Background:** Acute Hepatic Porphyria (AHP) is a rare genetic disease characterized by unpredictable life-threatening attacks. There is no reliable biochemical screening test for patients outside of an attack and diagnosis is delayed on average by 15 (!) years. AI screening systems can assist in detecting AHP patients, but validating such systems is challenging, due to the limited number of suspected candidates and/or success of recalling such candidates for testing. At the same time no study to date has highlighted human oversight of AI screening tools, while governing bodies and medical device regulations call for it to be allowed for clinical use. Our primary goal was to demonstrate the feasibility of an AI-based Human-in-the-Loop screening (HAI) approach and quantifying the added value by comparing the rate and number of clinically plausible cases found through it with the current Standard of Care (SOC).

**Methods:** This retrospective cohort study included data collected from 899,862 electronic health records (EHR) of patients who were treated at the University Hospital Salzburg (SALK) between December 2007 and December 2021. For our HAI approach we used an AI-tool for disease screening (Dx EHRs v2022.11) provided by Symptoma GmbH, that has been validated for Pompe Disease in a previous study. All historically suspected and diagnosed AHP cases retrieved from the collected data served as the reference standard representing the SOC. All suspected AHP cases were first triaged by generalist physicians (GP) without a specialization for AHP representing the “Humans in the Loop”. Specialized physicians (SP) determined the clinical plausibility of cases by reviewing the complete EHRs of the triaged cases. The primary outcome were the rates of clinically plausible cases (=precision) in the HAI and SOC cohorts and its sub-cohorts. Additionally, we investigated the differences in phenotypes in those cohorts. Historically diagnosed AHP cases were reviewed by SP for the reliability of their diagnosis.

**Findings:** Of a total of 899,862 EHRs, 191 EHRs were triaged into the HAI cohort and 107 filtered into the SOC cohort. 74 (38.74%) and 28 (27.72%) cases were deemed clinically plausible, for HAI and SOC respectively. Of those 74 clinically plausible cases in HAI, 46 were de-novo cases missed by SOC. The sub-analysis on the phenotypical features indicated that psychological and psychosomatic symptoms (Restlessness, Confusion, Anxiety, Depression, Mood swings, Palpitations) are significantly underrepresented within historically suspected AHP cases. As well were some common and subtle symptoms (Pain, Nausea, Vomiting, Fatigue). Among 16 historically diagnosed cases, four were reclassified as misdiagnosed, and seven lacked conclusive evaluation by current diagnostic standards. Notably, two new AHP cases were identified “incidentally” during the study, with a Poisson probability of 8.34% for this event to happen, suggesting this occurrence was unlikely to be random.

**Interpretation:** AHP is incredibly hard to diagnose and even already made AHP diagnoses are unreliable. Additionally, certain phenotypes are especially challenging to identify via the current standard of care. HAI managed to reach a higher precision compared to the SOC and found an additional 46 clinically plausible de-novo cases. Both showing feasibility and added value of HAI. “Incidentally” newly diagnosed AHP patients strongly suggest an increase in awareness through the AI screening project. All our findings suggest that HAI is a viable approach addressing the challenge of early diagnosis of AHP and its adherent issues. Prospective studies in a setting as real-time decision support at the point of care are warranted as a next step to implementing HAI as part of the new standard of care.

**Funding:** Alnylam Pharmaceuticals

**Research in context:** *Evidence before this study:* We systematically searched PubMed for articles published from database inception up to November 11, 2024, using the terms: ("Artificial Intelligence" OR "AI" OR "Machine Learning" OR "Deep Learning" OR "Neural Networks") AND ("Screening" OR "Diagnosis" OR "Detection") AND ("Rare Disease" OR "Orphan Disease" OR "Uncommon Condition" OR "Low Prevalence Disease" OR "Acute Hepatic Porphyria" OR "AHP" OR "Porphyria") AND ("Human-in-the-loop" OR "Human-assisted" OR "Human-centered" OR "Augmented intelligence" OR "Human-machine collaboration" OR "Human-computer interaction" OR "Hybrid intelligence" OR "Human-supervised" OR "Human oversight" OR "Collaborative AI" OR "Human-guided"). This search yielded no matches, suggesting that no dedicated studies investigating human-in-the loop AI approaches have been performed to date, neither for AHP, nor for rare diseases as a whole. We further adapted the search to look for AI screening approaches in general for Acute Hepatic Porphyria (AHP) specifically, by eliminating the search terms for rare diseases and human-in-the-loop. This search yielded 23 articles of which only two were actually related to the disease AHP. Those two studies aimed at testing AI screening systems for AHP patients but rather focused on successfully detecting AHP patients by using AI only, than comparing performances to the standard of care or highlighting human oversight. In those studies, no new cases could be found which appeared to be mainly due to the limited number of suspected candidates and/or success of recalling such candidates for testing which combined with the ultra-rare prevalence of AHP created unfavorable odds of finding de-novo cases. This stresses the incredible challenges when diagnosing AHP, but also in extension when validating new AI systems supporting such diagnosis. As such more validation in general, but much more direct comparison with the current state of the art is needed to ensure the effectiveness and safety of AI-driven diagnostic support systems for AHP. Hence, we did a retrospective cohort study aimed at evaluating the effectiveness of an AI-based Human-in-the-Loop screening (HAI) approach compared to the standard of care (SOC) in diagnosing AHP cases. Our study design addresses the challenges in validation and simultaneously puts a spotlight on human oversight, which should both contribute accelerating the adoption of AI screening to assist in rare disease diagnosis.

*Added value of this study:* To the best of our knowledge this is the first study to date to investigate the added value of a Human-in-the-Loop AI screening concept compared to the SOC for AHP or rare diseases. Explainable AI and particularly human oversight have been urgently demanded by governing bodies and regulators. The EU Artificial Intelligence Act even prescribes human oversight for any AI system designed to be used as a medical device in the European Union, which is founded in the desire to make AI applications as safe and reliable as possible. We present the results of our current study to highlight the feasibility and added value of the HAI concept as well as its potential as prospective real-time screening at the point of care. We further present findings of additional in-depth analyses delineating shortcomings regarding the diagnosis of AHP in the current SOC. Providing such evidence should serve as the bedrock to justify the considerable efforts necessary for implementation, but most of all, shorten the time to diagnosis effectively by using AI for rare disease patients suffering from diseases like AHP.

*Implications of all the available evidence:* Real-time decision support at the point of care has been identified as a pivotal lever to improve early diagnosis of AHP. At the same time eHealth infrastructure is being reformed and many initiatives around the world strive for higher levels of maturity. This paves the foundation for centrally orchestrated digital and AI-driven tools. Thus, scalable support systems exploiting this infrastructure are required and need to be validated. In our study, our proposed HAI screening approach showed a higher plausibility rate in suspected cases compared to the SOC and statistical testing could not find a statistically significant difference between the screening methods. Auxiliary findings in our study suggested that AI screening might create additional awareness at the point of care, which proves to be an effective agent in improving the diagnosis of AHP. Further, we found that there might be certain phenotypes that are harder to identify as AHP cases and that already made AHP diagnoses are unreliable. All our findings suggest that HAI is a viable approach addressing the challenge of early diagnosis and its adherent issues. They warrant further prospective studies in a setting as real-time decision support at the point of care while strongly advocating for an implementation into the clinical routine.

## Introduction

### Acute Hepatic Porphyria (AHP)

is a genetic disease characterized by life-threatening attacks. The estimated prevalence in the general population is about 9-10:1.000.000 and is therefore clearly classified as a "rare disease" which are defined as having a prevalence of less than 1:2.000. ^1,2^ AHP is caused by a defect in one of several enzymes of heme biosynthesis and symptoms are caused by the accumulation of toxic heme precursors, specifically aminolaevulinic acid (ALA).^3^ It consists of four different types depending on which metabolic step in the heme biosynthesis is affected: acute intermittent porphyria (AIP), variegate porphyria (VP), hereditary coproporphyria (HCP), and δ-ALA acid dehydratase porphyria (ADP) ^4,5^

### Obstacles in diagnosis

The clinical manifestations are highly variable and often non-specific, even within the four different types, and especially in adults with less fulminant and chronic courses.^6^ Further, in some cases (especially HCP and VP) confirmatory testing for AHP can only be considered reliable during an acute attack.^7^ Given all of that AHP is often confused for other misdiagnoses causing reported delays of an average of 15 (!) years from the onset of the first symptoms until the conclusive AHP diagnosis.^8^ Delayed diagnosis is a serious problem as commonly used medications can trigger or exacerbate acute attacks, and untreated attacks can result in permanent neurological damage or death.^9^ Also economically, untreated AHP patients present a considerable burden as on average they create 48 times (24k vs 0,5k/year) more healthcare costs compared to well controlled patients.^10^ However, Givosiran, a novel targeted orphan drug for AHP, that has been recently approved and shown to be distinctly more effective than the standard hemin therapy can cost up to 78% ($482,113; 95% CI=$373,638–$594,778) more in annual costs.^11–13^

### Screening for early diagnosis

In general, there is no reliable biochemical screening test for asymptomatic patients outside of an attack.^7,9,14^ This is further aggravated as testing of family members of an index patient revealed asymptomatic high excreters (ASHE), who have chronically elevated levels of ALA and PBG, in the absence of any symptoms.^7^ Although genetic markers are known for AHP, genetic testing should not be used as first-line testing for suspected AHP, but to confirm the diagnosis and type of AHP in patients with positive biochemical testing.^7,15^ The reason being that a) most carriers (estimated 1% phenotypic penetrance) of pathogenic variants in AHP genes never experience acute attacks in their lifetimes^15,16^, and b) the finding of a genetic variation of unknown significance (VUS) in a patient without biochemical evidence raises more questions than it answers.^7^ All this points at the pivotal requirement for an approach which is a) independent of biochemical or genetic testing, b) capable of taking the patients’ medical history into consideration, c) routing patients to the right specialist while being resource-efficient at scale. Particularly a rapid and easy-to-use point of care test is needed for a diagnosis in a timely manner. ^7^In summary there is a need on an individual, economic and general healthcare level for finding a viable approach to AHP screening for early diagnosis.

### Leveraging AI for diagnosis

Several AI-based approaches have been investigated for the purpose of supporting the diagnosis of rare diseases.^17,18^ Training data-driven AI models for rare diseases is particularly challenging as rare diseases are rare and thus training data is as well. Regarding AHP the challenge is even further aggravated as the rare, diagnosed cases that could be used for training are often not reliable true positives due to frequent misdiagnoses.^7^ Cohen et al. and Bhasuran et al. attempted to address AHP in their studies advancing the field of research in this area.^19,20^ Unfortunately, no new cases could be found which appeared to be mainly due to the limited number of appropriate candidates and/or success of recalling suspected cases for testing which combined with the ultra-rare prevalence of AHP created unfavorable odds of finding de-novo cases.^20,21^ As such more validation, specifically the direct comparison with the current state of the art, is still needed to ensure the effectiveness and safety of AI-driven diagnostic support systems for AHP.^22,23^ Providing such evidence will be crucial to justify the considerable efforts necessary for implementation.^24,25^ Most of all it should pave the way to eventually use AI to shorten the time to diagnosis effectively for rare disease patients suffering from diseases like AHP.^26^ In addition to that, explainable AI and particularly human oversight have been urgently demanded by governing bodies and regulators.^27,28^ The EU Artificial Intelligence Act prescribes human oversight for any AI system designed to be used as a medical product in the European Union, which is founded in the desire to make AI applications as safe and reliable as possible.^29^

We present the results of our current study to demonstrate the potential of an AI-based Human-in-the-Loop screening (HAI) approach which we are proposing as a prospective real-time screening at the point of care. We further present findings of additional in-depth analyses delineating shortcomings regarding the diagnosis of AHP in the current Standard of Care (SOC).

## Methods

### Study Design

This retrospective cohort study aimed at evaluating the effectiveness of an HAI approach compared to the SOC in detecting AHP cases. Data were collected from electronic health records (EHRs) of patients who were treated at the University Hospital Salzburg (SALK)^30^ between December 2007 and December 2021. There has never been any kind of AI-enhanced screening for AHP at SALK prior to our study.

### Outcomes

The primary outcome variable was the rate of clinically plausible cases (=precision) for AHP after the review of AHP specialist doctors. The primary independent variable was the type of screening approach (HAI vs. SOC). Secondary outcomes were the differences in symptomatic phenotypes of the clinically plausible portions of the two main cohorts (HAIp and SOCp). Secondary exploratory analyses evaluated the standalone performance of the AI tool by comparing the precision and number needed to screen of the AI alone cohort with the HAI (AI vs. HAI). Further a sub-analysis of the HAI sub-cohort HAI_de-novo_ compared the performance of HAI finding de-novo AHP cases with SOC.

### Data Collection

A total of 899.862 EHRs were collected. Each Electronic Health Record (EHR) contains an array of documents (specified in Table 1) related to an individual patient. All data was anonymized by the IT department of SALK prior to analysis. To account for historical AHP patients, the respective dataset was filtered using Regular Expression (REGEX) terms directly suggesting an AHP diagnosis or suspicion (i.e. ICD codes, disease name, etc.)(design of REGEX terms specified in the Supplementary Material) EHRs were categorized into two cohorts based on the type of screening: those were identified with (HAI and SOC). Further as HAI included historical AHP cases it was split into a sub-cohort including only de-novo AHP cases (HAI_de-novo_) for better interpretability regarding added value and contrast to SOC. In addition to the “screened” cohorts, a “Background” cohort was generated from the remaining patients. Selection was biased such that only EHRs were selected for the “Background” cohort which showed at least one feature related to AHP (exemplary list of features specified in the Supplementary Material). Inclusion and exclusion criteria for all EHRs were only for them to be technically not corrupted.

**Table 1.** Array of Document Types contained in the electronic health records (EHRs)

| Documents |
| --- |
| 1. Entries on allergies |
| 2. Anamnesis/Patient History |
| 3. Discharge letter |
| 4. Specialist Findings |
| 5. Entries on departments visited and case ID |
| 6. Doctor's notes |
| 7. ICD - codes |
| 8. Procedure codes |
| 9. Vitals and other measurements |
| 10. Laboratory result values |
| 11. List of medications |
| 12. Surgical reports |
| 13. Administrative patient profile |
| 14. Nurses' notes |
| 15. Radiology reports |
| 16. Entries on social habits/risk factors |
| 17. Wound reports |

### Pre-processing

To standardize the representation of EHR data, we mapped the data to one data model, adhering to the Fast Healthcare Interoperability Resources (FHIR) standard. This mapping ensured the data to be stored in a uniform manner, facilitating more streamlined processing and analysis downstream. As part of data cleaning, we removed duplicated records and excluded EHRs which were not unambiguously assigned to a single case or individual.

### Screening

The proposed screening approach followed a Human-in-the-Loop concept. An AI-tool for disease screening (Dx EHRs v2022.11) was provided by Symptoma GmbH^31^, a medical software manufacturer specialized in medical software and diagnostic decision support systems.^32–37^ The tool generates a list of EHRs which have an increased probability to suffer from a specific rare disease - in our current study AHP. It has been validated for Pompe Disease in a previous study and is under active development.^38^ Prior to applying to the collected EHRs the performance has been tested in-silico on a synthetic dataset to ensure the feasibility for AHP. (Results and Methodology in the Supplementary Material) After EHRs were flagged by the tool as suspected cases for AHP, generalist physicians (GP) without a specialization for AHP triaged the results for further review by specialized physicians (SP). GP categorized the EHRs into *“*Diagnosed*,”* “Suspected,” “Reduced Suspicion,” and “Rejected” (definitions in Table 2) to facilitate further review, while “Rejected” EHRs were excluded for the review by SP. Two SP, consultants with particular expertise in the field of genetic metabolic diseases (GS, EA), reviewed the results and rated the clinical plausibility following a hierarchical consensus principle, where a decision can be overruled by the more senior physician (EA), if no consensus could be reached. The SP assigned the labels “Definite,” “Probable,” “Possible,” “Inconclusive,” and “Unlikely” (definitions in Table 3). EHRs labeled either “Rejected” by the GP or “Unlikely” by the SP were considered as “not clinically plausible” for further analysis. All others were considered “Clinically Plausible”. To ensure “Rejected” cases by GP equaled “Unlikely” cases according to SP, 44 “Rejected” cases (10% of all flagged EHRs) were presented to SP for double verification. This methodology and the associated labels were selected to minimize workloads for physicians and to optimally align with clinical decision-making processes. (Figure 2 shows the process flow in detail) Deceased cases were excluded from analyses.

**Table 2.** Classifications by Generalist Physicians (GP)

| Label | Explanation |
| --- | --- |
| **Diagnosed** | Medical review found a documented diagnosis for Acute Hepatic Porphyrria (AHP) |
| **Suspected** | Medical review found supporting clinical evidence for AHP |
| **Reduced suspicion** | Medical review found conflicting clinical evidence for AHP |
| **Rejected** | This patient will be rejected due to the medical review result overriding the AI result |

**Table 3.** Classifications by Specialist Physicians (SP)

| Label | Explanation |
| --- | --- |
| **Definite** | Acute Hepatic Porphyrria (AHP) is the top differential diagnosis for the given case |
| **Probable** | AHP is in the top ten of differential diagnoses for the given case |
| **Possible** | AHP is a possible differential diagnosis for the given case |
| **Inconclusive** | There is insufficient evidence speaking for or against AHP as a differential diagnosis for the given case |
| **Unlikely** | AHP is not among the differential diagnoses for the given case |

### Phenotypical analysis

For the sub-analysis of the differences in phenotypes, we defined two sub-cohorts consisting of all EHRs deemed clinically plausible by SP in the respective main cohorts HAI (HAIp) and SOC (SOCp). To avoid introducing biases of historically diagnosed and treated cases, we removed those from SOCp. To avoid introducing noise of historical AHP cases, we removed those from HAIp and only included de-novo identified cases. Further, we compare those sub-cohorts to a Background cohort. For the comparison 88 phenotypical features in eight main categories (Hematological, Dermatological, Gastrointestinal, Neurological, Psychiatric, Cardiopulmonary, Pain-Related, Miscellaneous) related to AHP were extracted (full list of features is specified in the Supplementary Material).

### Additional Findings

To investigate the patient journey of historically diagnosed cases in more detail SP reviewed their EHRs and summarized findings regarding their work up status. Findings were summarized into three categories: “Confirmed Diagnosis”, “Misdiagnosis”, “Further Work-Up required” (definitions in Table 4). Information on de-novo AHP cases at SALK was aggregated from anonymous data collected through unstructured interviews with SP (EA, GS), which are employed consultant physicians at SALK.

**Table 4.** Assessment categories for the patient journeys of historically diagnosed cases.

| Label | Explanation |
| --- | --- |
| **Confirmed Diagnosis** | Acute Hepatic Porphyria (AHP) has been correctly diagnosed according to the state-of-the-art diagnostic criteria |
| **Misdiagnosis** | AHP is a misdiagnosis according to the state-of-the-art diagnostic criteria |
| **Further Work-Up required** | AHP cannot be confirmed based on the documented work-up |

### Statistical Analysis

Data are presented using descriptive statistics. Further analysis using the chi-square test for independence was performed to compare the rate of clinically plausible cases for AHP between the cohorts HAI_de-novo_ and SOC. The null hypothesis was for the plausibility rate in HAI_de-novo_ to be equal to SOC. The alternative hypothesis was that HAI_de-novo_ has a different plausibility rate compared to SOC. A contingency table was constructed with the rows representing the screening approaches (HAI_de-novo_ and SOC) and the columns representing the clinical plausibility rating (not/clinically plausible). The chi-square statistic was calculated, and the corresponding p-value was obtained. For the analysis of the differences in phenotypes the p-values were calculated by a two-tailed Fisher’s exact test and corrected for multiple testing by the Benjamini-Hochberg method. The probability of finding new AHP cases in a defined timeframe based on a historic expectancy value was calculated using the Poisson probability mass function. P-values less than 0,05 were considered statistically significant. The confidence intervals were calculated assuming binomial distributions and a confidence level of 95%. Chi-square test and descriptive statistics were calculated using jamovi (version 2.5.4), an open-source statistical software. Phenotype analyses and probability calculations were performed using SciPy (version 1.14.0), an open-source software for mathematics, science, and engineering.

### Ethical Considerations

The study was conducted in accordance with the Declaration of Helsinki and approved by the Institutional Review Board of the Federal State of Salzburg (EK Nr. 1006/2023). Informed consent was waived due to the retrospective nature of the study. Patient confidentiality was maintained by anonymizing all data before analysis.

## Results

### Flow of results

In Figure 1 we present the complete flow of results. Of a total of 899,862 EHRs, 546 EHRs were flagged by the AI for suspicion of AHP, of which 44 cases had been deceased and thus excluded for further review. Of the remaining 502 flagged EHRs, 191 EHRs were triaged by GP into the Human-in-the-loop Screening cohort (HAI) for further review by SP. Of those 6 were found to be already deceased and of the remaining 185 EHRs, 74 were rated clinically plausible by SP. At the same time 107 EHRs with historical suspicion for or diagnosis of AHP were identified using REGEX and thus included in the standard of care cohort (SOC), six were already deceased and thus excluded from further analysis, of the 101 remaining EHRs 28 were rated plausible by SP. Table 6 indicates the numbers of not/clinically plausible cases in those 3 cohorts.

**Figure 1.**
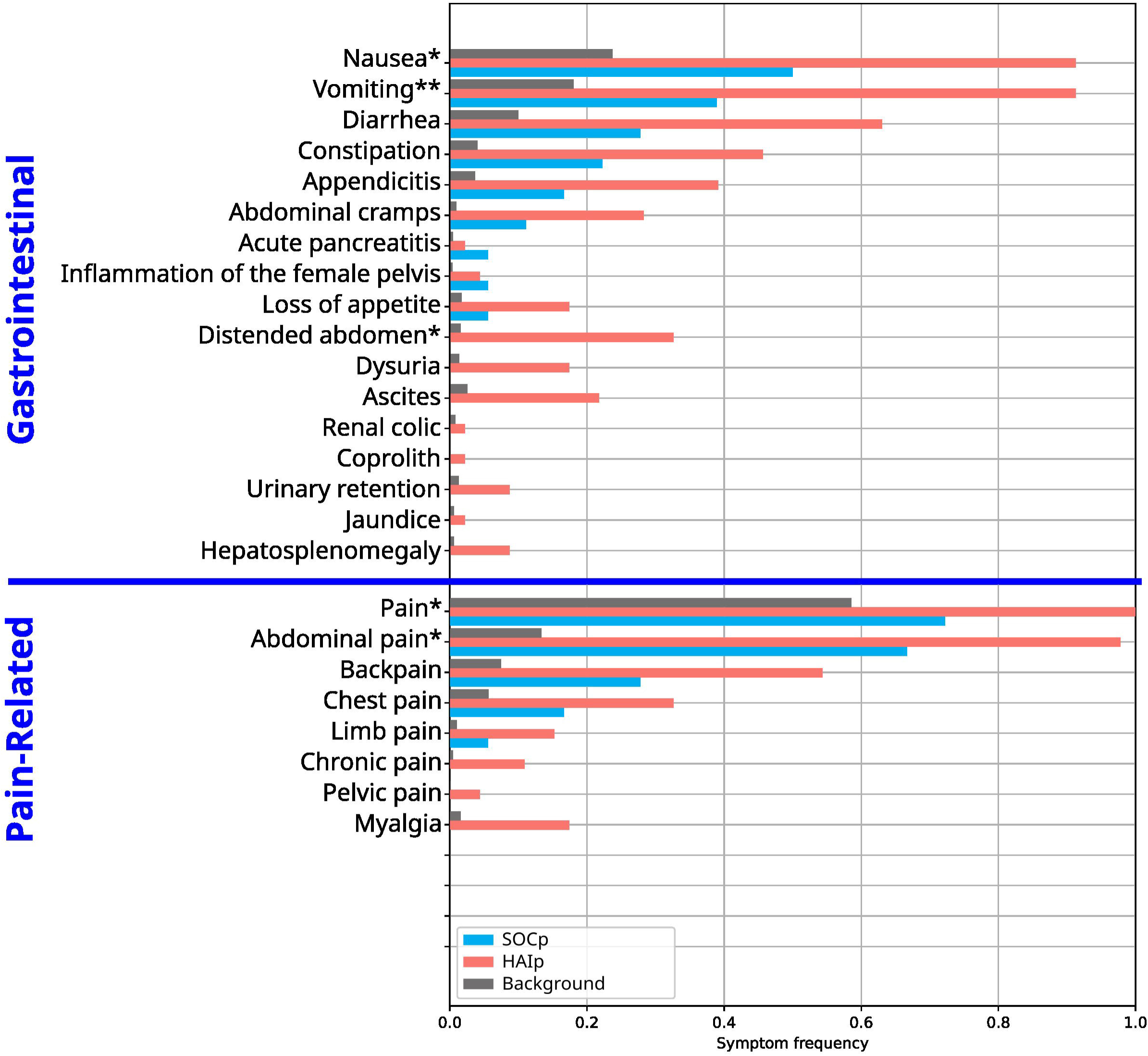
Complete flow of results for AI/HAI and SOC through the 2 stage appraisal by GP and SP. Results are colour coded: Orange= “Suspected” cases by GP; Yellow= “Reduced Suspicion” cases by GP; Red= “Rejected/Unlikely” cases by GP/SP; Grey=Deceased cases; Light Green= “Clinically Plausible” Cases (“Definite,” “Probable,” “Possible,” “Inconclusive” cases by SP

**Figure 2.**
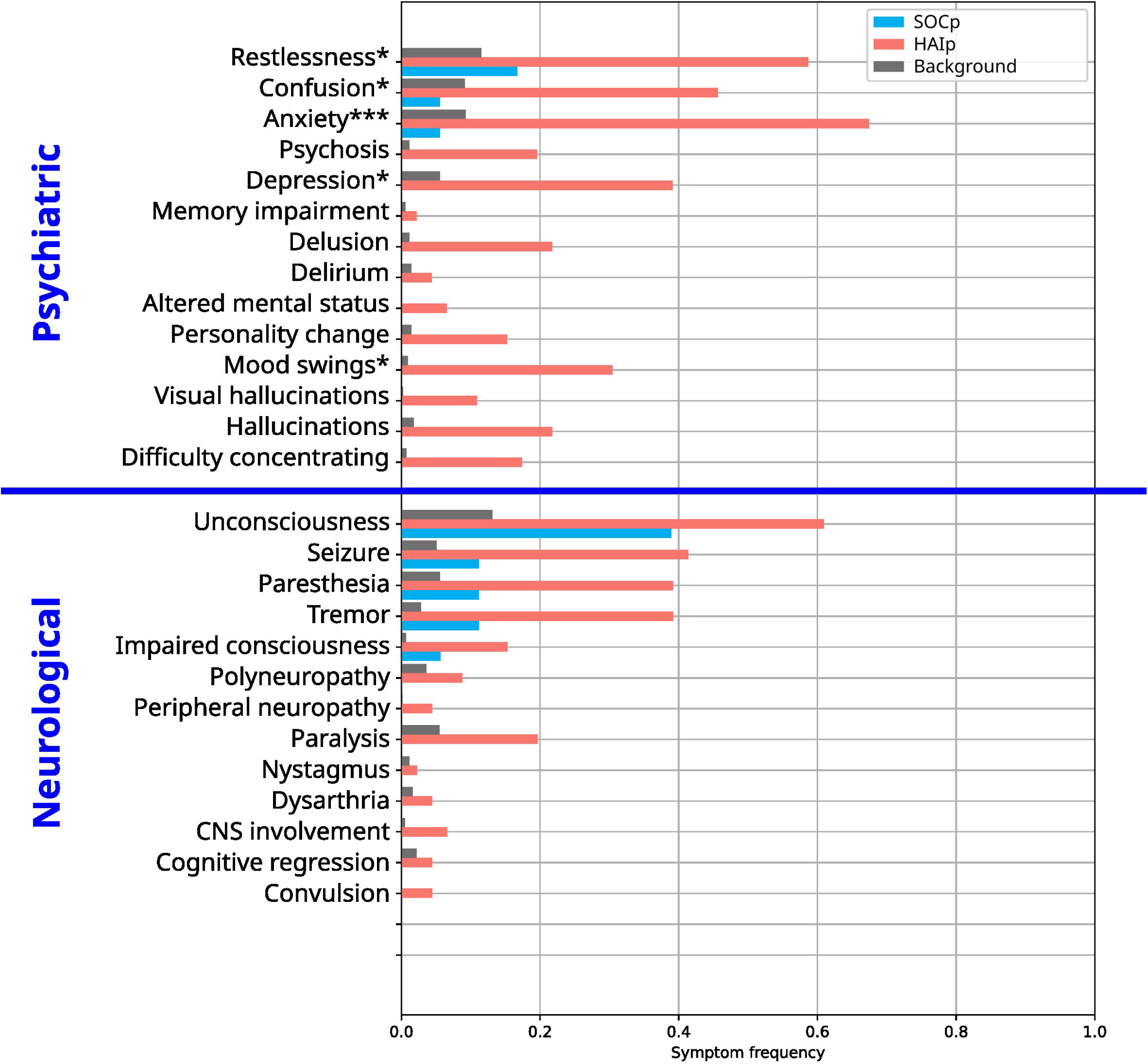
Step-by-step process overview of our proposed Human-in-the-loop AI Screening

**Figure 3.**
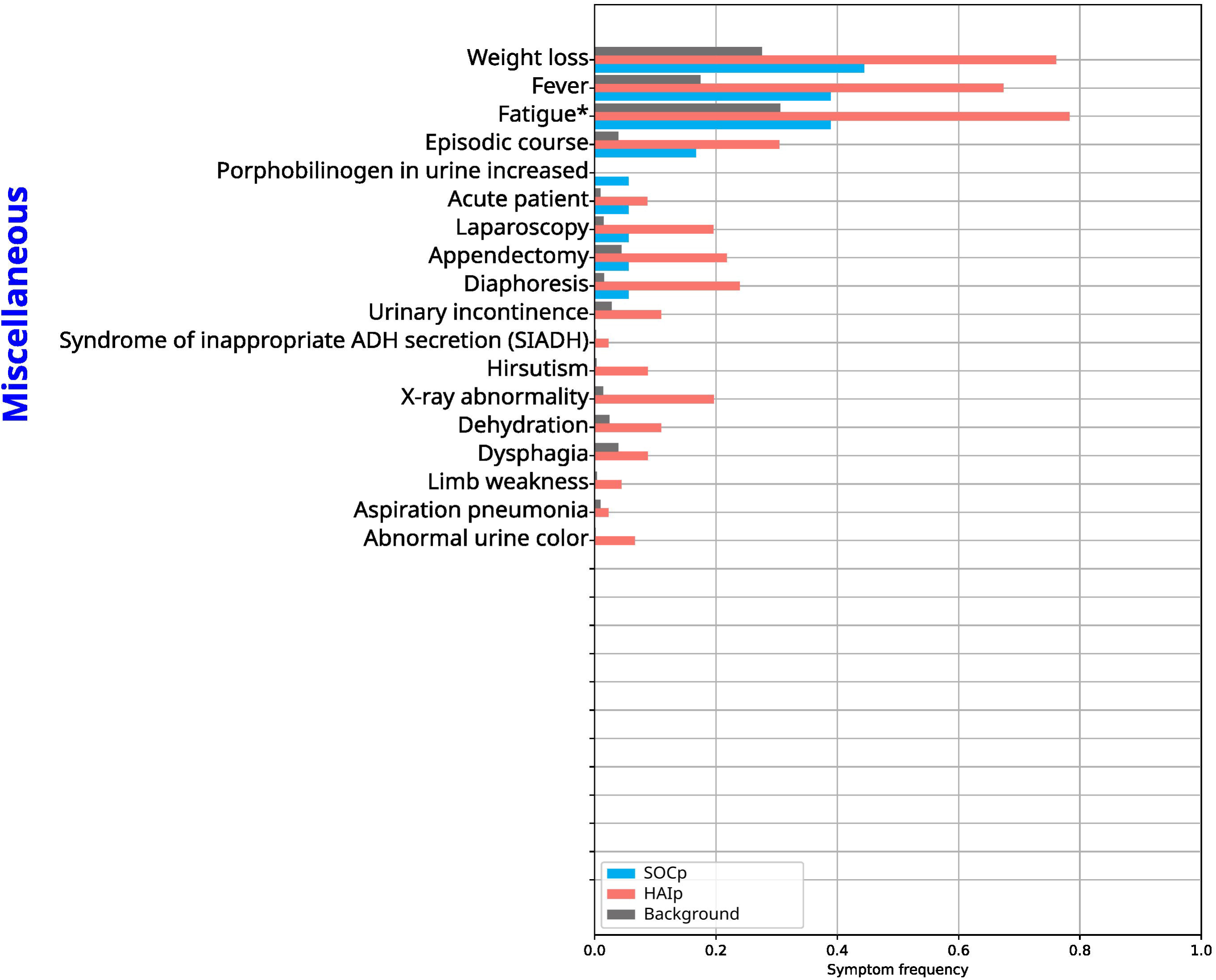
Our proposed Human-in-the-loop AI Screening as a centrally installed diagnostic support system covering all potential harbors of clinical first contact for potential cases (Primary and Secondary Care). The dotted lines show the secure server environments where data can be analyzed and stored safely (Green=Hospital Facility; Cyan= Health Insurance infrastructure)

### Demographic characteristics

All cohorts but the SOC cohort (54.5% male) form predominantly female populations. SOC is also the oldest population (mean=52.4 years, SD=23.1) accompanied by the highest age at first admission (mean=39.1 years, SD= 22.7). The HAI_de-novo_ cohort showed the on average longest treatment duration (mean=11.3 years, SD= 4.4). Table 5 summarizes the descriptive statistics on the age, age at first admission, treatment duration and sex in each cohort.

**Table 5.**
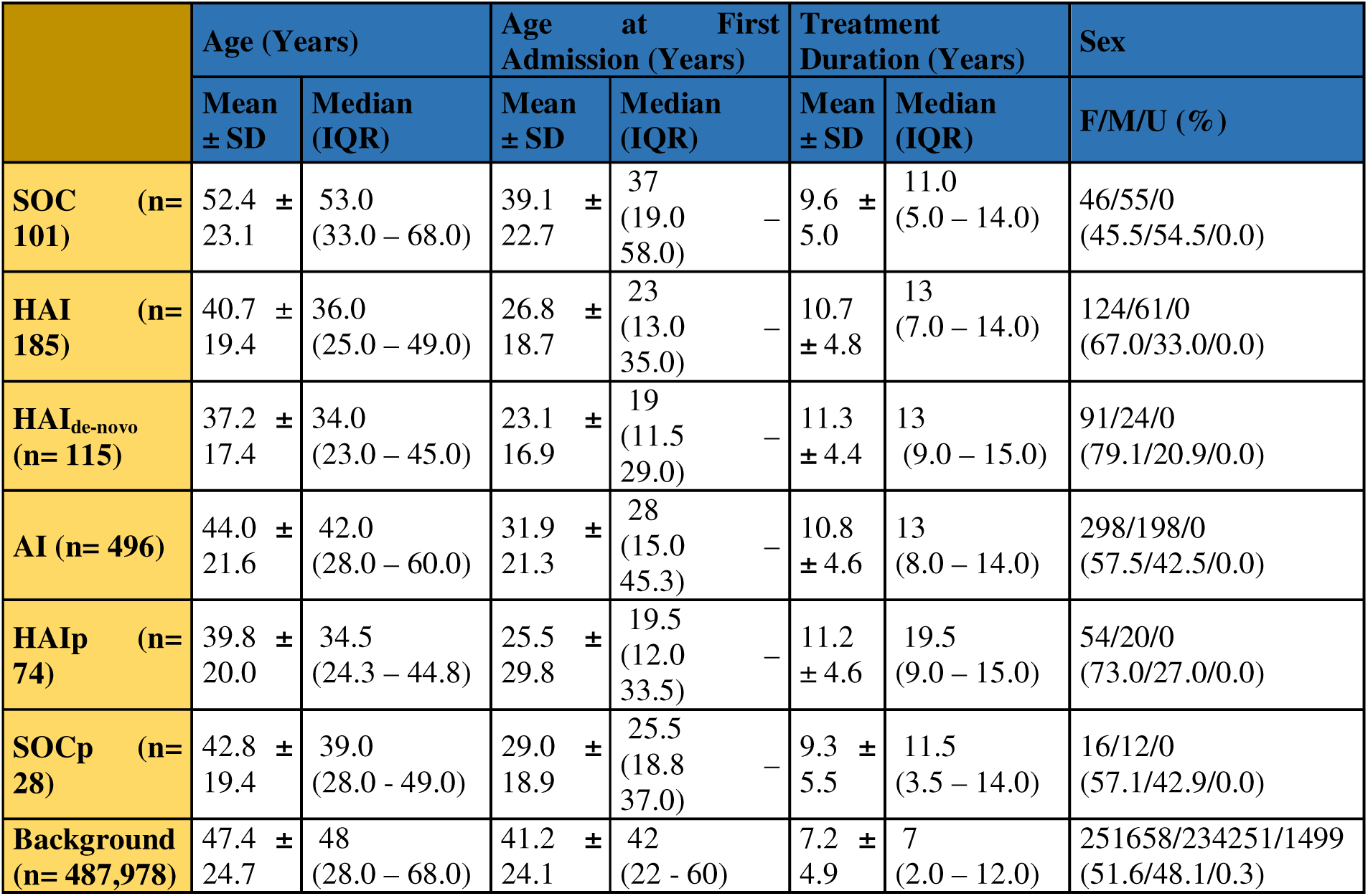
Descriptive statistics of the demographic characteristics in each cohort. The treatment duration was calculated as the time difference between the age at first and last recorded admission. Data are n (%) for categorical variables and mean ± SD and median (IQR) for continuous variables. (SD= Standard Deviation; IQR= Interquartile Range; F=Female; M=Male; U=Unkown)

**Table 6.** Number of clinically plausible AHP cases for cohorts SOC, HAI and HAI_de-novo_.

|  | Clinically Plausible | Not Clinically Plausible | SUM |
| --- | --- | --- | --- |
| **Human-in-the-Loop (HAI)** | 74 | 111 | 185 |
| **Human-in-the-Loop excl. historical cases (HAI<sub>de-novo</sub>)** | 46 | 69 | 115 |
| **Standard of Care (SOC)** | 28 | 73 | 101 |

### Human-in-the-Loop efficiency

546 EHRs were flagged by the AI for suspicion of AHP of which 50 cases had been deceased already and thus excluded for further analysis. Assuming an AI only screening (AI) approach, a precision of 14.91% would have been reached for finding clinically plausible cases vs. 40.00% with a human in the loop. SP would have had to review 496 (95% CI: 452.36 – 539.64) EHRs compared to 185 (95% CI: 163.89-206.11) in the approaches without human (AI) and with human in the loop (HAI), respectively. This equals a factor of 2.68 (95% CI: 2.19 – 3.29) more workload for specialist doctors without human in the loop.

### Phenotypical differences in historical and new cases

We performed a sub-analysis on the phenotypical features and compared the differences between cohorts (HAIp, SOCp, Background). The outcome is presented in Figure 4-Figure *7*, which show the relative frequency (RF) in each cohort. In Table 7 we present the features that were found to be significantly different between HAIp and SOCp.

**Figure 4.**
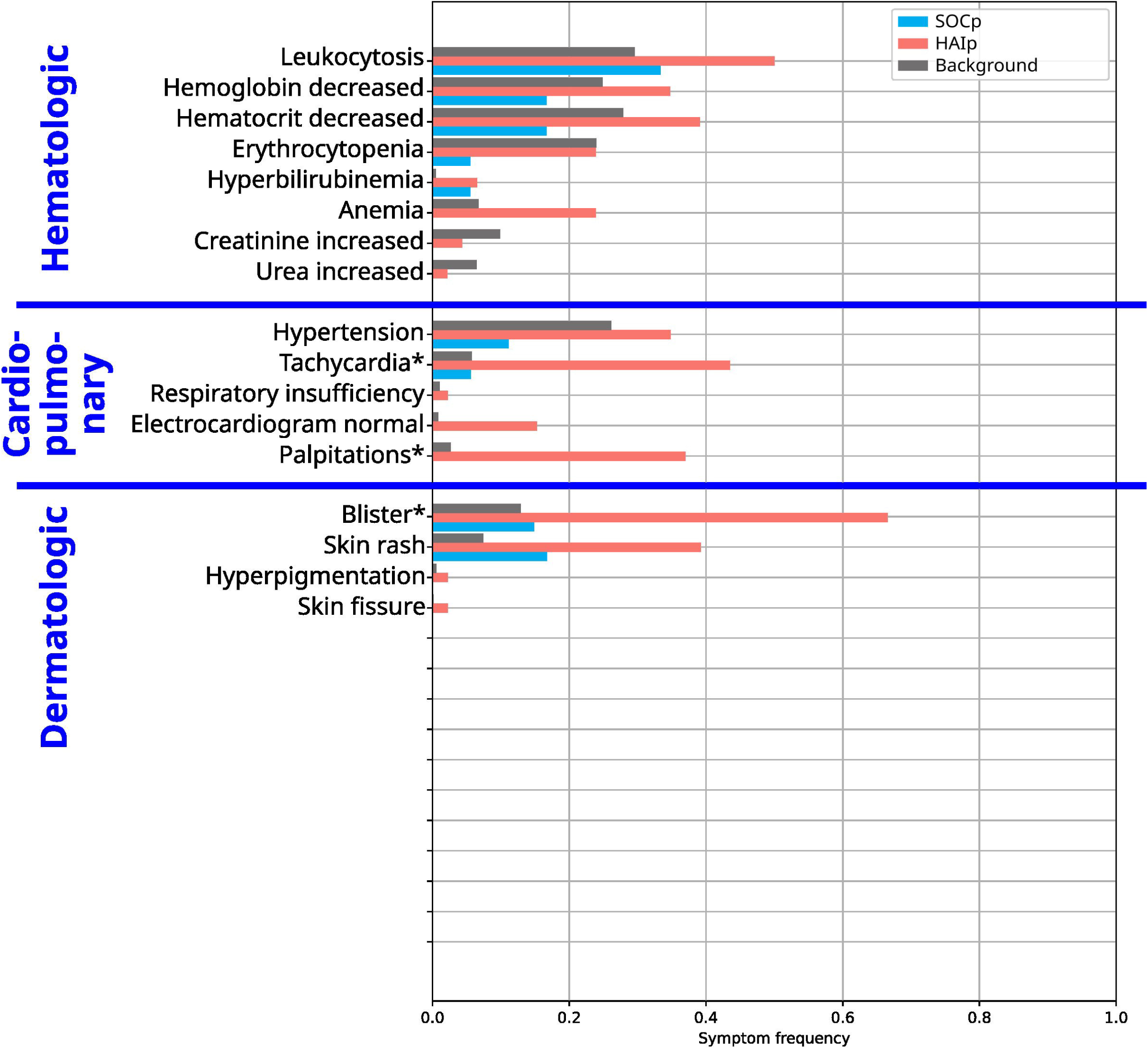
Bar chart showing the relative frequency (RF) of gastrointestinal and pain-related AHP symptoms in each cohort (HAIp n=46, SOCp=18, Background n= 487,978). Significance of the difference between these cohorts are indicated with one, two, and three asterisks which correspond to a p-value less than 0.05, 0.01, and 0.001 respectively.

**Figure 5.**
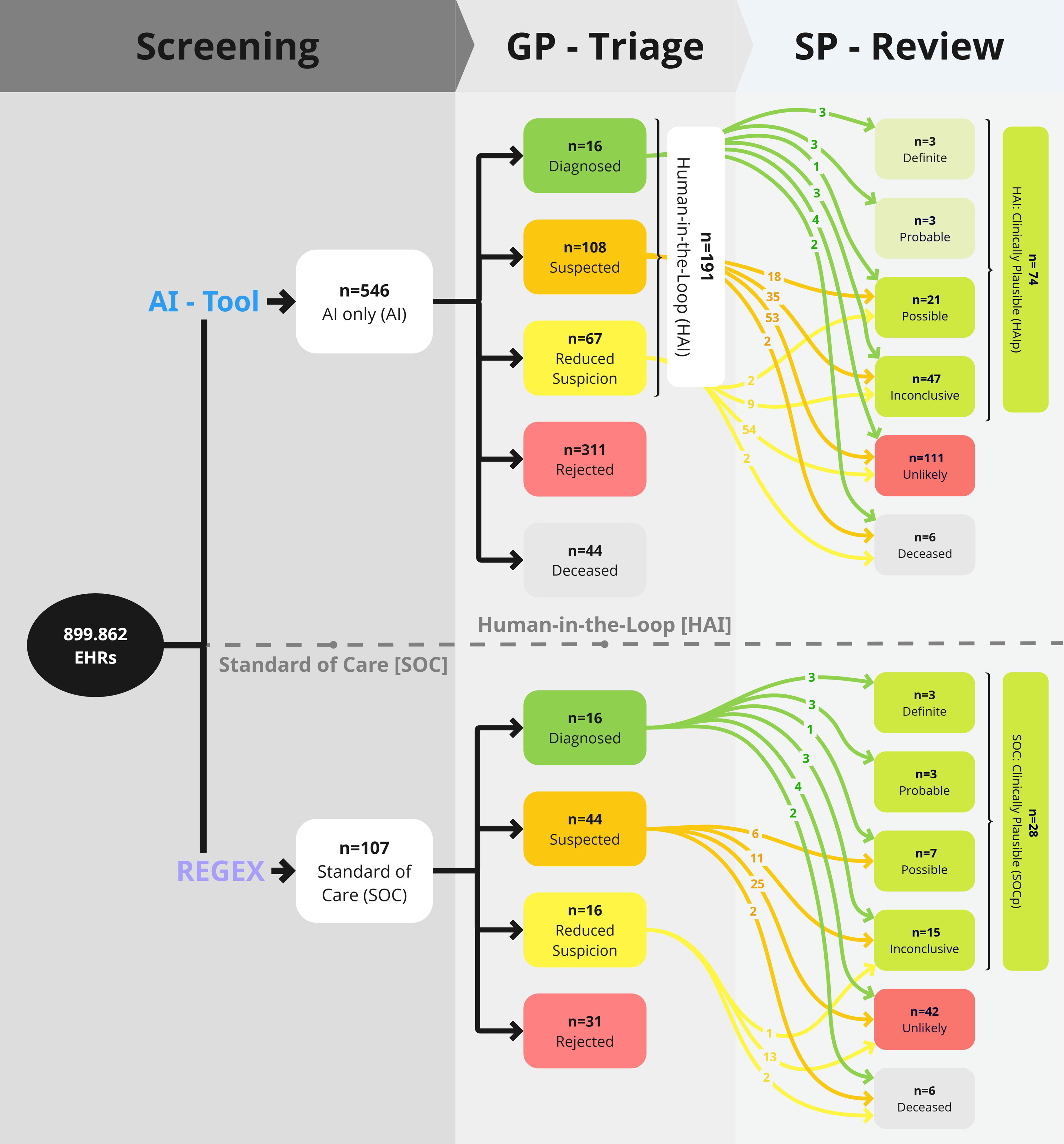
Bar chart showing the relative frequency (RF) of hematologic, cardiopulmonary and dermatologic AHP symptoms in each cohort (HAIp n=46, SOCp=18, Background n= 487,978). Significance of the difference between these cohorts are indicated with one, two, and three asterisks

**Figure 6.**
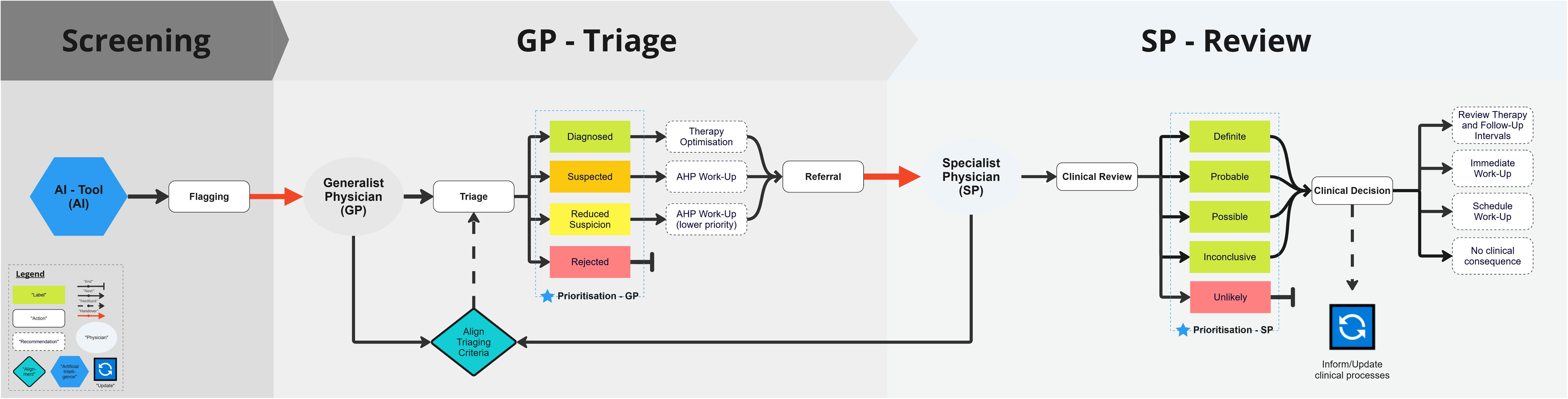
Bar chart showing the relative frequency (RF) of psychiatric and neurological AHP symptoms in each cohort (HAIp n=46, SOCp=18, Background n= 487,978). Significance of the difference between these cohorts are indicated with one, two, and three asterisks

**Figure 7.**
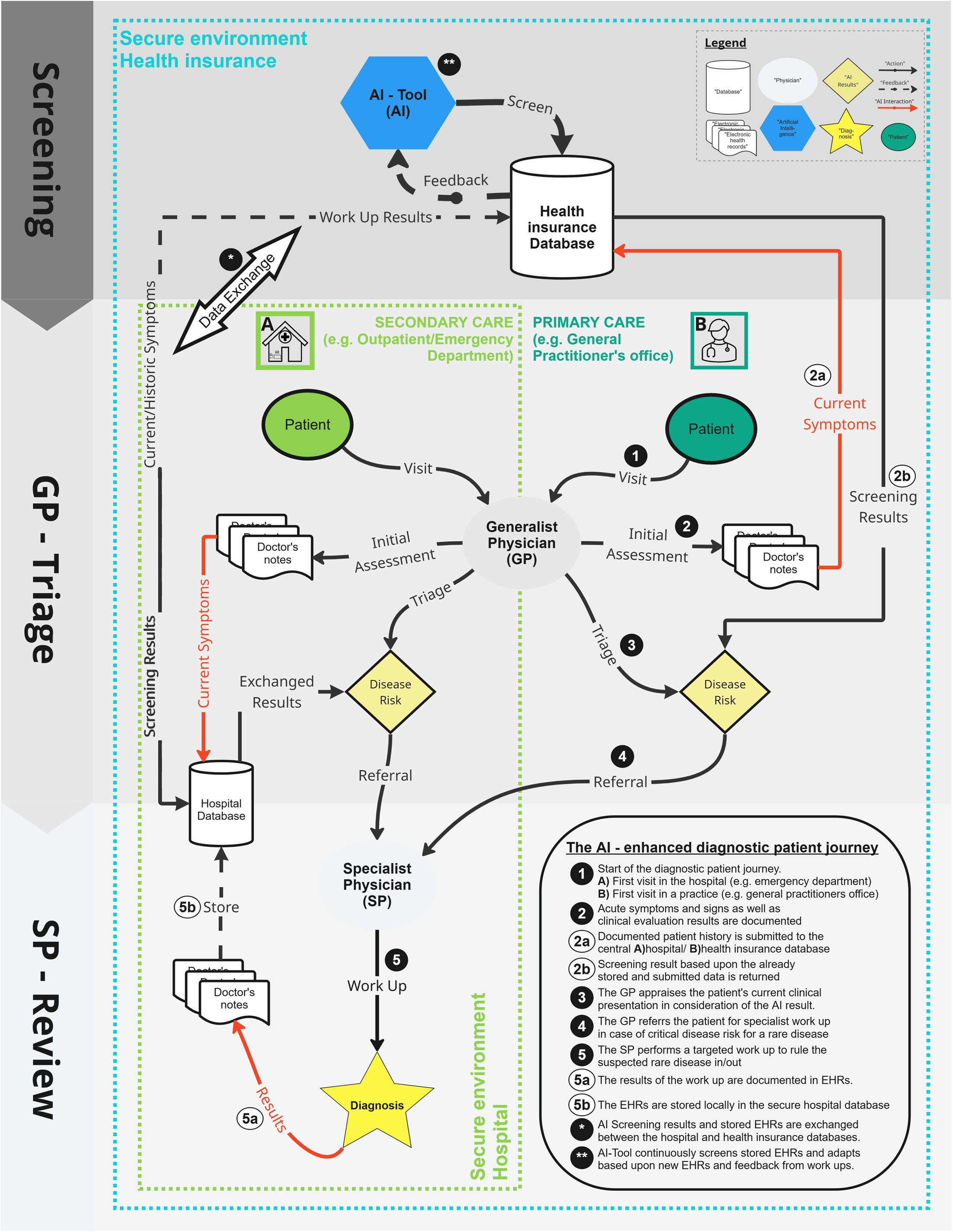
Bar chart showing the relative frequency (RF) of miscellaneous AHP symptoms in each cohort (HAIp n=46, SOCp=18, Background n= 487,978). Significance of the difference between these cohorts are indicated with one, two, and three asterisks

**Table 7.** All significantly different phenotypical features between HAIp and SOCp (RF=Relative Frequency; OR= Odds Ratio)

|  | Feature | RF<sup>HAIP</sup> | RF<sup>SOCp</sup> | OR | p-value (corrected) |
| --- | --- | --- | --- | --- | --- |
| **Dermatological** | Blister | 0.500 | 0.111 | 8 | 0.036 |
| **Gastrointestinal** | Nausea | 0.913 | 0.5 | 10.50 | 0.019 |
|  | Vomiting | 0.913 | 0.389 | 16.50 | 0.002 |
| **Miscellaneous** | Fatigue | 0.783 | 0.389 | 5.66 | 0.041 |
| **Pain-Related** | Pain | 1 | 0.722 | inf | 0.019 |
|  | Abdominal Pain | 0.978 | 0.667 | 22.50 | 0.019 |
| **Psychiatric** | Restlessness | 0.587 | 0.167 | 7.11 | 0.036 |
|  | Confusion | 0.457 | 0.056 | 14.28 | 0.030 |
|  | Anxiety | 0.674 | 0.056 | 35.13 | 0.001 |
|  | Depression | 0.391 | 0 | inf | 0.019 |
|  | Mood swings | 0.304 | 0 | inf | 0.041 |
| **Cardiopulmonary** | Tachycardia | 0.435 | 0.556 | 13.08 | 0.031 |
|  | Palpitations | 0.37 | 0 | inf | 0.019 |

#### Additional Findings

##### Off-track cases

After the review of the EHRs of historically diagnosed cases by SP, four misdiagnoses have been discovered. Further, seven of 16 cases had been deemed to be diagnosed historically but not worked up conclusively. Table 8 summarizes the assessment results of all historically diagnosed cases.

**Table 8.** Summary of Specialist Physicians (SP) assessment results of the patient journeys of historically diagnosed cases (* 1 of 7 cases could be recalled and confirmed as correct AHP diagnosis later)

|  | Patient Journey Stage | Cases (%) |
| --- | --- | --- |
| **Historically Diagnosed (n=16)** | Confirmed Diagnosis | 3 (18.8) |
|  | Misdiagnosis | 4 (25.0) |
|  | Further Work-Up required | 7* (43.8) |
|  | Deceased | 2 (12.5) |

##### Awareness effect

In parallel to our study two new cases of AHP were diagnosed by AHP disease specialists (all in 2023) with no prior documented visit at SALK. Based on the expectancy value of four historically discovered and confirmed cases in 15 years from 2007 to 2021, the chance of finding two new AHP cases in two years is an estimated 8,34% using the Poisson probability mass function.

## Discussion

### Clinical Need

Looking at the results of our current study, it suggests that almost two thirds of all clinically plausible AHP cases might be missed by the current Standard of Care (HAI_de-novo_ clinically plausible: n=46 vs. SOC clinically plausible: n=28). However, even if diagnosed with AHP, the diagnosis was not correct or conclusive in most cases (Confirmed diagnosis: n= 4 (25%)). Those findings highlight the urgent clinical and research need to take action towards improving the diagnosis of AHP, which has also been identified by the European Union Committee of Experts on rare diseases (EUCERD).^39,40^ Improving diagnosis for AHP patients addresses several game-changing pain points around AHP patients, the biggest being the diagnostic delay of on average 15 years from the onset of symptoms until AHP diagnosis.^8^ Furthermore, untreated patients experience a high health burden in terms of acute attacks, chronic symptoms, and long-term complications.^41^

However, the critical first step for diagnosis is the suspicion for AHP at the point of care and consequent referral to an expert for specialized work up. In our study the mean age and age at first admission is the highest in the SOC cohort (SOC: 52.4 years and 39.1 years versus HAI_de-novo_: 37.2 years and 23.1 years), revealing a considerable potential to catch patients earlier. At the same time, it shows that AI can help with that. This is further highlighted by the natural course of the disease beginning most commonly between the twenties and forties.^42,43^ The SOC cohort showed also an unexpected male dominated distribution in sex (54.5% male) compared to all other cohorts, although women are predominantly affected by AHP. ^43,44^ This may point at another systemic shortcoming where HAI_de-novo_ showed (79.1% female) it can support addressing it.

### Systemic support

Generalist physicians in i.e. practices, outpatient and emergency departments need to be supported in the initial triaging process which is not well established yet. ^26,45^. Using an AI-tool to assist with the identification of suspicious AHP cases can improve early diagnosis and support addressing aforementioned issues, while automating the bulk of the workload. Figure 3 shows our proposed Human-in-the-loop setup as a centrally installed diagnostic support system covering all potential harbors of first clinical contact for potential cases (Primary and secondary care). In some European countries the infrastructure has already reached or is close to reaching this level of maturity (e.g. Austria: ELGA, Denmark: Kliniske kvalitetsdatabaser) while others having concrete plans to get there soon (e.g. Germany: ePA).^46–48^ However, having a system setup directly in the secure environment of a hospital can be a lower threshold option for the near-term as an intermediate step towards scaling the diagnostic support.

### Human-in-the-Loop approach

The goal of this study was to demonstrate the feasibility and added value of a practical screening approach to improve the early diagnosis of AHP patients. Schreiweis et al. found in their systematic review of facilitators for novel eHealth solutions that when reaching the level of implementation the main facilitating factors found were ease of use, integration into care, and user-friendliness, all pointing at the crucial point of avoiding to disrupt existing processes^25^. Thus, we chose to pilot an approach, which would integrate into existing diagnostic pathways, while minimizing the disruption of existing clinical processes. We aimed at a setup which would both reduce the workload for the small number of existing specialized physicians (SP), while maintaining human oversight of the AI results through “generalist physicians” (GP) which are more readily available within existing healthcare structures. As such, we mimic the organic patient journey, starting at the general practitioner’s office, or an outpatient department and consequent referral to specialists when suspicion for AHP arises. Having those two stages (GP, SP) also enables a more refined triaging approach. (Figure 2: Dashed line “Prioritisation - GP/SP”) SP can align with GP on specific hard and soft criteria for triaging the urgency for referral (Figure 2: Box “Align Triage Criteria”) such as age or sex, as AHP rarely develops before puberty and women are more frequently affected.^42–44^ SP can triage according to the level of suspicion(Figure 2: Dashed line “Prioritisation - SP”) and clinical need for further intervention, while informing existing clinical routines (i.e. (re-)evaluating AHP Patient follow-up procedures) and diagnostic protocols (i.e. update state-of-the-art testing guidelines). In Figure 2 we present the process flow showing the different stages along the AI-enhanced diagnostic pathway.

### Clinical Utility

Despite increasing evidence of AI and eHealth solutions being powerful tools for healthcare applications, a lack of studies demonstrating a clinical benefit slow down the uptake of such solutions.^17,49^ By comparing the performance of our proposed approach to the state-of-the-art we are clearly demonstrating the potential clinical utility and attempting to address at least in part the frequently reported gap between tested accuracy and clinical utility.^23^. Looking at the results, the performance of this setup (Precision_HAI_ = 40.00%) proves to be competitive with a clear edge over the standard of care (Precision_SOC_ = 27.72%). However, this includes historical cases which have been flagged by the AI as well. This makes the direct comparison harder as historical cases are prone to be found by the AI as they contain more and also explicit data features pointing at AHP. To investigate the difference further we created a sub-cohort only containing de-novo suspected AHP cases. This way an added value can be contrasted even further by evaluating the precision of HAI when discovering de-novo cases. The chi-square test failed to prove a statistically significant difference between Precision_HAIde-novo_ and Precision_SOC_, which is generally challenging to achieve with rare disease studies but also considering the pre-test probability being decreased as historically suspected and diagnosed EHRs are excluded from the HAI_de-novo_ cohort. ^50,51^ However, this also means that HAI is able to identify de-novo cases with a comparable effectiveness as the SOC, which is an important finding underlining its capability to provide added value in practice. Interestingly, while historical AHP cases could be considered “must-not-miss” cases, also the misdiagnosed cases were flagged by the AI. This points at the authority of the information provided in the EHRs serving as the ground truth and at the limitation of the support AI can provide. Part of it can be explained by the purpose of the AI tool, which is to screen, and as such to favour sensitivity over specificity. But also, EHR documentation is burdened by subjectivity and an already made diagnosis can bias the documented information swaying both AI and GP to agree with the historical judgement. In summary, the performance for the HAI_de-novo_ cohort was superior as well (Precision_SOC_ = 27.72% vs. Precision_HAIde-novo_ = 40.00%) and no significant difference could be found to the SOC (p = 0.058), however caution should be given when re-appraising historical cases, as those are also hard for a HAI screening to debunk.

### AI pertinence

In general, but especially with rare diseases, it is of utmost importance to increase the pre-test probability to lower manual workloads and optimize the predictive power of diagnostic testing. Already using AI alone in our study, we increased the pre-test probability to find clinically plausible cases by more than ∼4800 – fold. That is from roughly 0.003% (= 28 plausible cases in 899,862 EHRs manually worked up in the clinical routine) to 14.91% (74 plausible cases in 496 EHRs suggested by AI). With our HAI approach SP had to review 2.5 cases to find 1 clinically plausible case which is another ∼2.7 times more efficient than AI only. Due to the low prevalence of rare diseases (AHP prevalence: 9-10:1,000,000), the number of patients screened is the most relevant variable for success.^1^ Thus, resource-efficient scalability in addition to accuracy are key metrics for any screening approach. An AI-based approach scales with increasing resource efficiency for every additional patient included for screening as the required resources stay roughly the same independent of the number of patients. This indirect proportional behavior marks a pivotal advantage for the screening of rare diseases. In comparison screening 899.862 cases manually would theoretically take one physician roughly 45.000 days or 123 years straight, while in our study assisted with AI the number of cases requiring human screening was reduced to 496 which would take one physician roughly 25 days to complete.^52^ An AI-based approach also presents further advantages particularly catering to challenges in the diagnosis of rare disease such as exploiting existing retrospective EHRs. This enables a complete and conclusive analysis considering all historical records of each individual reaching back in time which addresses the issue of premature closure, a common reason for misdiagnosis.^53,54^

### Awareness

An additional effect of an AI-based screening appears to be that it effectively creates awareness at the point of care. Healthcare professionals who are seeing, diagnosing, and treating potential AHP cases are directly reminded of the existence and presentations of AHP in the clinical routine. When considering the expectancy value of four confirmed correctly diagnosed cases in a timeframe of 15 years from 2007 to 2021 (Table 4), the chance of finding two de-novo AHP cases in two years is only 8.34%. This suggests an indirect effect of our current study, which was unexpected, but nevertheless had a huge impact for the affected AHP patients. While beyond expectation in our study, Kosch et al. showed that expecting benefits from using an AI can already improve human performance.^55^ Further it has been observed in other studies that even sham AI technology can have robust placebo effects, leading to participants gather information more quickly, and make them more alert.^56,57^ No matter the exact mechanism behind the seemingly increased awareness for AHP, the beneficiaries are certainly the patients. Further, heightened awareness in the clinical routine can also benefit existing patients as closer monitoring of existing AHP patients helps ensuring optimized prophylactic treatment, which does not only benefit patients directly but has also shown to have economic benefits by reducing required resources.^58^

### Reasons for misdiagnosis

Psychological and psychosomatic (Restlessness, Confusion, Anxiety, Depression, Mood swings, Palpitations) symptoms are significantly underrepresented within historically suspected AHP cases pointing at a systemic issue with linking them to AHP. However, psychological symptoms are as relevant and common as other key symptoms like pain-related ones are.^6,59^ Additionally, more unspecific and subtle symptoms (Pain, Nausea, Vomiting, Fatigue) highlight the importance of linking common symptoms to ultra-rare diseases such as AHP, which is known to be an issue.^7,60^ This is especially critical as those are some of the most frequently reported chronic AHP symptoms. ^61^ One dermatological sign (Blister) is underrepresented within historically suspected cases which could be due to the fact that cutaneous manifestations are generally seen in VP only and rarely in other types of AHP. However, in cases of severe acute attacks, elevated porphyrin levels can cause blisters and fluid-filled bullae in VP and HCP stressing their general significance, but less so as standalone symptoms.^62,63^ The investigation results of the diagnostic journeys of historically diagnosed AHP patients prove again how difficult a reliable diagnosis of AHP is and moreover how much expert matter knowledge is required to perform a conclusive AHP work up. ^7,14^ Thapar et al. even suggest in a recent review of obstacles in AHP diagnosis that clinicians should never assume that a historically documented AHP diagnosis is accurate and laboratory results that led to the initial diagnosis must be reviewed - if not available – redone.^7^ Our results underpin those findings repeatedly, unfortunately.

### Outlook

Real-time decision support at the point of care has been identified as a pivotal lever to improve early diagnosis of AHP^7,14^. Some retrospective screening approaches have been attempted, however recall of patients proved to be challenging (response rate 5%-12%)^20^. At the same time eHealth infrastructure is being reformed and many initiatives around the world strive for higher levels of maturity.^64–67^ This may improve communication pathways with patients for better recall and also pave the foundation for centrally orchestrated digital and AI-driven tools. Meanwhile, further local and scalable efforts are required to help find patients and validate support systems retrospectively and prospectively. Thus, adding to the bedrock of evidence and pushing the envelope towards eventually implementing real time screening on a large scale.

### Limitations

### Clinical Plausibility

Clinical plausibility is only a proxy outcome for the actual diagnosis and the validation is dependent on what has been documented. However, all cases had been diligently cared for as part of the clinical routine, and it has also to be assumed that the pre-test probability for AHP of the de-novo investigated population (n = 899,755 EHRs) has already been reduced. Further, the patient recall of retrospective cases for confirmatory disease testing has been proven to be highly challenging.^20,21^ As such we opted to compare clinical plausibility rates between HAI and SOC. This primary outcome variable is not dependent on the success of patient recall ensuring a reliable outcome measure to compare the performances. In any case, clinical plausibility remains highly relevant at the point of care, as it determines the critical crossroads in every rare disease patient’s diagnostic journey of being referred to the right specialist.^26,45^

**Generalizability** is limited as only one AI system has been applied with this HAI concept and the data from one hospital analyzed. While the outcomes of our current study might have limited validity in representing performances beyond our local setup, a similar independent study in Oregon, USA indicated similar performances (“AI” 22 of 90 = 24,4% vs “Standard of Care” 24 of 97 = 24,7%) for a different healthcare region across the globe.^19^ This indicates a level of general validity to some extent. Although EHRs from a secondary care facility do not perfectly represent all patients at the beginning of their patient journey, their advantage lies within the quantity and quality of documentation. Further, comparing EHRs across primary and secondary care facilities would introduce additional biases through different software, documentation styles and potentially highly skewed sample sizes, burdening any comparison. Moreover, rare disease patients have been characterized as a cohort with highly diverse largely non-linear diagnostic journeys which might be better represented within a secondary care facility, which includes a variety of potential points of encounter (i.e. emergency, outpatient, inpatient departments etc.).^26,53,68,69^

### Standard of Care

Although a retrospective analysis of the historical management of AHP in the past 15 years might not perfectly depict the current clinical practices today, it does allow for a fair representation of the “Standard of Care”. The “Standard of Care” in medicine is not only defined by the existing methods and products, but also by the experience and professional expertise applied to the individual case. It is informed by empirical evidence and as novel and improved tools become available, the lower threshold for the state of the art moves upwards.^70^ New standards must first be proven to be consistently superior, easily integrated into existing workflows and financially viable for the respective healthcare provider, for it to be implemented.^27^ As such there is a certain bandwidth of products, methods and practices which form the “Standard of Care”.

## Conclusion

**In summary**, our proposed HAI screening approach showed higher plausibility rates compared to the existing standard of care and discovered an additional 46 clinically plausible de-novo cases. Auxiliary findings in our study showed that HAI screening identifies de-novo cases with a comparable precision as the SOC and suggest it might create additional awareness at the point of care. Substantiating HAI screening further as an effective agent in improving the diagnosis of AHP. Moreover, we found that there might be certain phenotypes that are harder to identify as AHP cases and that already made AHP diagnoses are often unreliable. All our findings highlight that the diagnosis of AHP remains a challenge for the standard of care and they suggest that an AI-based human-in-the-loop screening is a viable approach to address this challenge and its adherent issues. They warrant further prospective studies in a setting as real-time decision support at the point of care while strongly advocating for an implementation into the clinical routine.

## Data Availability

The source data used for this study comprise electronic health record (EHR) data containing protected health information under European GDPR from patients under care at University Hospital Salzburg (SALK). Due to legal agreements with the providing institution (SALK), the anonymized datasets analyzed during the study are not publicly available. Aggregated data of anonymized datasets in tabular format can be made available upon reasonable request to the corresponding author, subject to institutional approval. Requests to access these datasets should be directed at.

Full details regarding the implementation and setup of the AI tool (Dx EHRs v2022.11) are available from Symptoma GmbH upon request. Investigators with expertise in the field should be able to reproduce the described methods on their own data to validate the results of this study.

## Contributors

SL and JN contributed to the conception and design of the study. SL, KB, MK, VP and GS compiled the database and implemented the methodology. SL and TL performed the data analysis. JN was responsible for funding acquisition. SL was responsible for project administration. SL wrote the first manuscript draft. SL, JN and GS revised the manuscript critically. All authors had access to the data used in the study, contributed to the article and approved the submitted version.

## Acknowledgments

This work is part of the PhD thesis of the first author SL (Department of Internal Medicine, Paracelsus Medical University, Salzburg, Austria). We are grateful for the contributions to the data preparation and analysis from Nicolas Munsch and Rafael Weingartner-Ortner. We want to thank Prim. Univ.-Prof. Dr. Elmar Aigner (EA) for his support in the initiation of and his clinical guidance during the study. This study was funded by Alnylam Pharmaceuticals. Alnylam Pharmaceuticals was not involved in the study design, collection, analysis, interpretation of data, the writing of this article or the decision to submit it for publication.

