## Supplementary Material for "Unmet Needs in Acute Hepatic Porphyria Diagnosis: A Comparative Big Data Analysis of an AI-based Human-in-the-Loop Screening Versus Standard of Care"

### In-silico performance

**Rationale for the in-silico study.** This in-silico study was performed at the planning stage of the project. As such it served the purpose of gauging the technical feasibility in preparation of the project.
An in-silico study design was chosen which is reproducible AND resource light, as any efforts invested need to be pertinent to the purpose. (Project preparation vs. Project implementation).
Further the validation of the AI tool is done in stages to increase confidence in the predicted performance. This is also in line with the EU ethics guidelines of creating “Trustworthy AI”.^[1]^

The initial hypothesis for this in-silico study was, “Is Symptoma’s AI-tool (AI) able to detect a disease like Acute Hepatic Porphyria (AHP) with very vague symptoms and differentiate against other diseases which overlap in their presentations?”

**Methodology.** To demonstrate the capability of the AI to identify AHP we performed an in-silico study and analyzed a total of n = 7112 medical test cases. The different sets and sources of these cases are described as follows:

To avoid selection bias when creating the test set symptom frequencies for acute and chronic AHP were taken from an independent study by Gouya et al.^[2]^, who characterized a cohort of AHP (n=112) patients over the course of 6-12 months.

AHP cases: We make use of the symptom list from Gouya et al. to construct synthetic cases yielding a total number of 6000 synthetic cases (3000 for the acute and 3000 for the chronic presentation of AHP). A case is created by adding each symptom from AHP with a probability equal to the symptom frequency. For example, if the frequency of Nausea is 40%, the probability of adding Nausea for a case is 0.4.
Control cases: A total of n = 1112 cases were sourced from the British Medical Journal (BMJ) and transcribed by a medical doctor into sets of symptoms, both negative and positive, alongside other risk factors, the patient’s age, and the patient’s sex when available.
The cases cover a diverse range of causes, including patients suffering rib fractures, rabies, or metastatic cancer. The number of symptoms and keywords per case ranges from 1 to 33 (median = 8) including complex terms such as “right true vocal cord is immobile”.
A subset of the most pertinent cases was created by taking only those which included symptoms associated with AHP. Namely, cases which reported any of the symptoms from Figure 1.
Under this constraint, 515 out of the 1112 BMJ cases were selected to compare with the acute presentation of AHP and 405 with the chronic. These subsets represent cases with higher similarity to AHP presentation and, hence, an increased chance of misdiagnoses.

**Results.** We consider a patient as AHP positive if their symptoms result in AHP to be returned in the first 30 diagnoses listed by the AI. AHP returned below this, or not at all, results in a patient being classified as AHP negative. This method of evaluation reflects the same one used in the most comprehensive review of symptom checkers to date^[3]^ as well as our prior publications.^[4]^^[5]^^[6]^

It provides an evaluation of accuracy with regards to AHP under the criteria used for the assessment of general diagnoses. Under this definition, the AI classifies nearly all 3000 acute AHP synthetic cases correctly as AHP cases (Figure 2: 97% accuracy, 97% f-1 score, 100% sensitivity, 94% specificity, 95% precision). Without further differentiation of the algorithms for the chronic presentation of AHP, the AI still classifies most of the 3000 chronic synthetic AHP cases correctly as AHP cases (Figure 2: 76% accuracy, 71% f-1 score, 59% sensitivity, 93% specificity, 90% precision). The interesting aspect to this analysis is that acute cases are based on 16.59 and the chronic on 2.33 symptoms on average, which is determined by the symptom frequencies given in Figure 1. For the chronic presentation many asymptomatic cases (0 symptoms) were generated as to the frequencies given. In subsequent analyses with at least 1, 2, 3, 4, and 5 symptoms the accuracy improves from 79 to 95% respectively even for the chronic cases (Figure 3). Already with 3 symptoms available the AI was able to infer the risk for AHP with +90% precision. This is particularly important for settings where only limited data on the patient is available, i.e. first visit(s) of the patient.

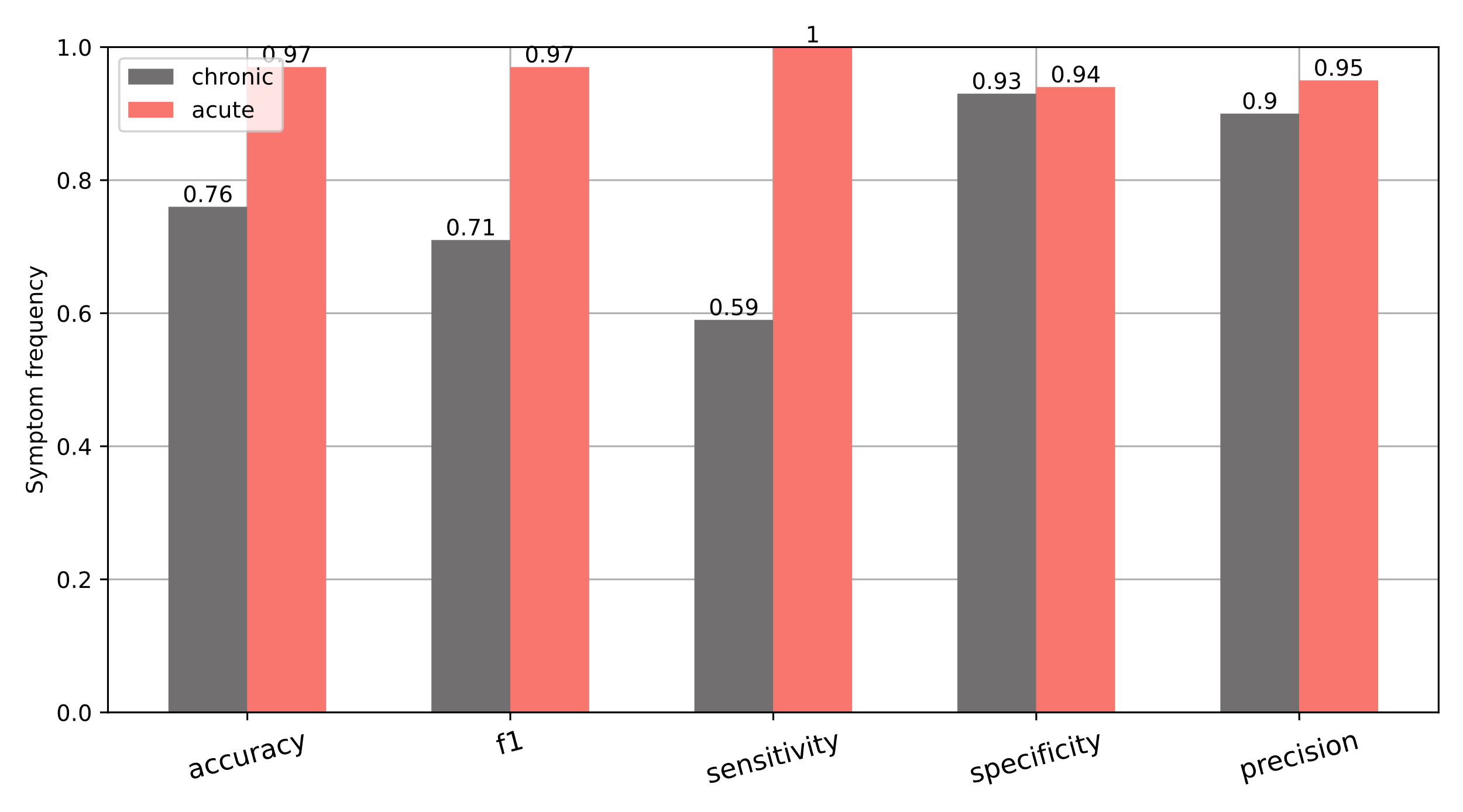

Figure 8 Performance metrics of the AI for acute and chronic, synthetic AHP cases

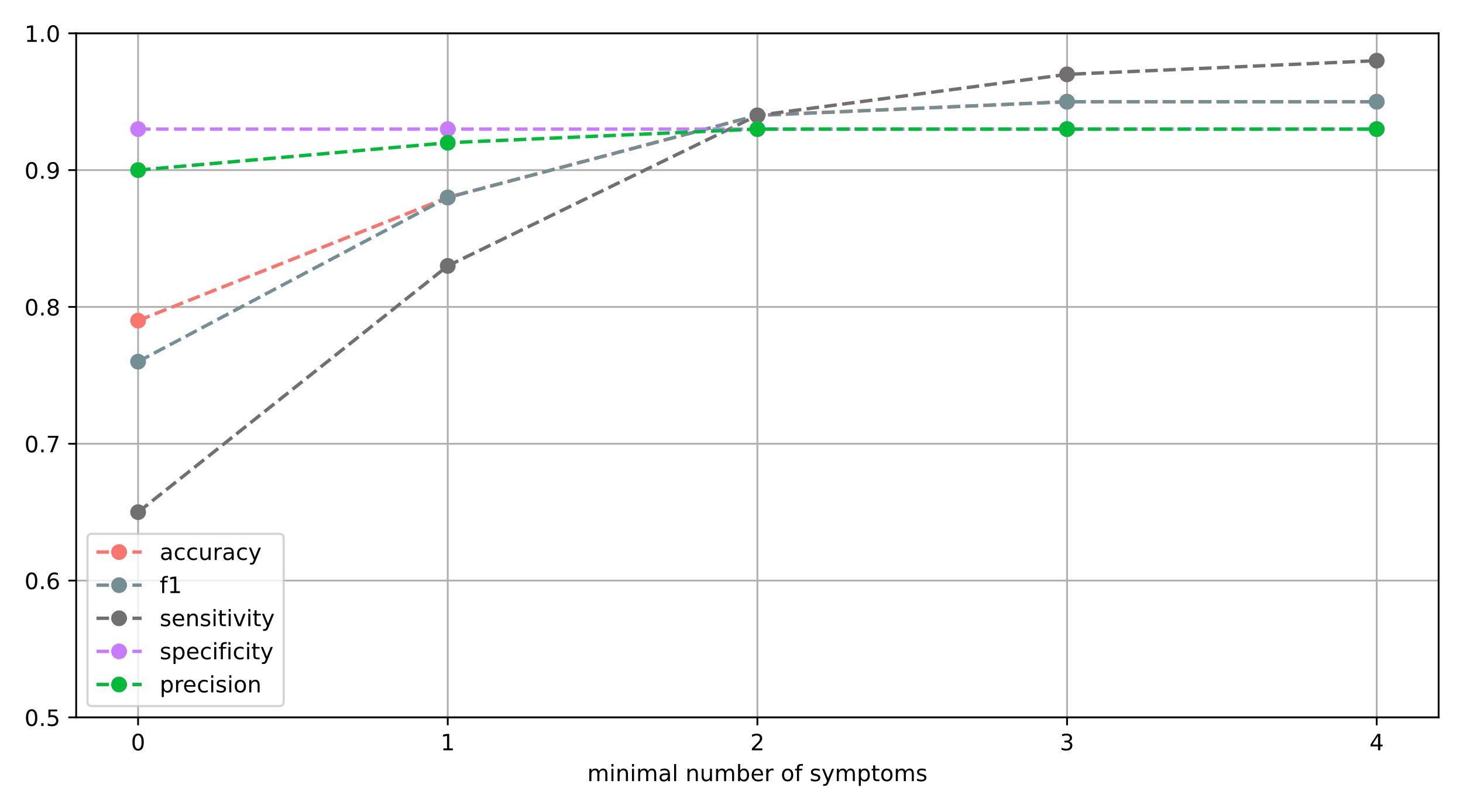

Figure 9 Accuracy of the AI over number of minimum symptoms per case for the chronic, synthetic AHP cases

**Discussion.** When assessing performance results from in-silico studies it is important to understand the gaps between in-silico and in-vivo conditions. In this case our in-silico data was generated from averaged symptom frequencies (Gouya et al.) and standardized clinical vignettes (“BMJ cases”). However, it was indicative to see promising performance metrics for both acute and chronic presentations of AHP. Despite the fact that those two clinical presentations were virtually generated from natural histories. As such the purpose was to test the general capability of the AI to differentiate permutations of AHP cases in different disease phases when encountered in everyday practice. However, confounders like documentation styles, incomplete documentation or atypical AHP presentations can only be accounted for with limited confidence. As such in-silico testing will inherently carry biases when comparing to in vivo test settings. This rightfully must be accounted for when extrapolating performance metrics to real world settings. However, the purpose was to estimate the feasibility of the AI to differentiate AHP against other diseases which overlap in their presentations as part of the preparatory work for the actual study. The study itself allows for a more accurate evaluation of the capability in a real-world setting using real-world data.

### Design of REGEX terms to identify historical AHP cases

To design REGEX terms which would ensure finding historical AHP patients we included all possible names and abbreviations for AHP (i.e. acute hepatic porphyria (AHP), acute intermittent porphyria (AIP), variegate porphyria (VP), hereditary coproporphyria (HCP), and δ-ALA acid dehydratase porphyria (ADP) etc.). Further we included diagnostic tests or pathognomonic signs for AHP (i.e. positive Watson Schwartz test, elevated urinary delta-aminolevulinic acid, increased urinary porphobilinogen, etc.)

The following shows examples for how the REGEX terms were created:

- r"Akute (Porphyrien hepatisch|Porphyrie hepatisch)",
- r"Akute hepatische Porphyrie",
- r"Hepatische (Porphyrien akut|Porphyrie akut)",
- r"Positiver Watson Schwartz Test",
- r"(Delta(-| )Aminol(ä|ae|a|e)vulins(ä|ae|a|e)ure(-| )Dehydratase|Porphobilinogen(-| )Synthase|Porphyrie aufgrund eine.? Delta(-| )Aminolevulinat(-| )Dehydratase)(-| )?(Mangel|Defizit)?",

### List of AHP features to characterize phenotypical differences

**Hematological**

Leukozytose

Hämoglobin erniedrigt

Hämatokrit erniedrigt

Erythrozytopenie

Hyperbilirubinämie

Anämie

Kreatinin erhöht

Harnstoff erhöht

**Gastrointestinal**

Nausea

Erbrechen

Diarrhoe

Konstipation

Appendizitis

Abdominale Krämpfe

Akute Pankreatitis

Entzündung des weiblichen Beckens

Appetitverlust

Aufgeblähtes Abdomen

Dysurie

Aszites

Nierenkolik

Koprolith

Harnretention

Ikterus

Hepatosplenomegalie

**Neurological**

Bewusstlosigkeit

Krampfanfall

Parästhesie

Tremor

Bewusstseinsstörung

Polyneuropathie

Periphere Neuropathie

Lähmung

Nystagmus

Dysarthrie

ZNS-Beteiligung

Kognitive Regression

Konvulsion

**Miscellaneous**

Gewichtsverlust

Fieber

Ermüdung

Episodischer Verlauf

Porphobilinogen im Urin erhöht

Akutpatient

Laparoskopie

Appendektomie

Diaphorese

Porphyria cutanea tarda

Harninkontinenz

Syndrom der inadäquaten ADH-Sekretion

Hirsutismus

Röntgen abnormal

Dehydratation

Dysphagie

Gliedmaßenschwäche

Aspirationspneumonie

Urinfarbe abnormal

**Pain-Related**

Schmerz

Bauchschmerzen

Rückenschmerz

Brustschmerz

Gliederschmerzen

Chronischer Schmerz

Beckenschmerzen

Myalgie

**Psychiatric**

Rastlosigkeit

Verwirrtheit

Angst

Psychose

Depression

Gedächtnisstörung

Wahn

Delir

Geisteszustand verändert

Persönlichkeitsveränderung

Stimmungsschwankungen

Optische Halluzinationen

Halluzinationen

Konzentrationsschwierigkeiten

**Cardiopulmonary**

Hypertonie

Tachykardie

Respiratorische Insuffizienz

Elektrokardiogramm normal

Palpitationen

**Dermatological**

Blase

Hautausschlag

Hyperpigmentation

Hautriss

1. https://digital-strategy.ec.europa.eu/en/library/ethics-guidelines-trustworthy-ai
2. Gouya L, Ventura P, Balwani M, Bissell DM, Rees DC, Stölzel U, et al. EXPLORE: A Prospective, Multinational, Natural History Study of Patients with Acute Hepatic Porphyria with Recurrent Attacks. Hepatology. 2020 May;71(5):1546-1558.
3. Semigran HL, Linder JA, Gidengil C, Mehrotra A. Evaluation of symptom checkers for self diagnosis and triage: audit study. BMJ. 2015 Jul 8;351:h3480. doi: 10.1136/bmj.h3480. PMID: 26157077; PMCID: PMC4496786.
4. Munsch N, Martin A, Gruarin S, Nateqi J, Abdarahmane I, Weingartner-Ortner R, Knapp B. Diagnostic Accuracy of Web-Based COVID-19 Symptom Checkers: Comparison Study. J Med Internet Res. 2020 Oct 6;22(10):e21299. doi: 10.2196/21299. PMID: 33001828; PMCID: PMC7541039.
5. Martin A, Nateqi J, Gruarin S, Munsch N, Abdarahmane I, Zobel M, Knapp B. An artificial intelligence-based first-line defence against COVID-19: digitally screening citizens for risks via a chatbot. Sci Rep. 2020 Nov 4;10(1):19012. doi: 10.1038/s41598-020-75912-x. PMID: 33149198; PMCID: PMC7643065.
6. Nateqi J, Lin S, Krobath H, Gruarin S, Lutz T, Dvorak T, Gruschina A, Ortner R. Vom Symptom zur Diagnose – Tauglichkeit von Symptom-Checkern : Update aus Sicht der HNO [From symptom to diagnosis-symptom checkers re-evaluated : Are symptom checkers finally sufficient and accurate to use? An update from the ENT perspective]. HNO. 2019 May;67(5):334-342. German. doi: 10.1007/s00106-019-0666-y. PMID: 30993374.
